# Proteomic Insights into Lp(a) Cardiovascular Mechanisms: A Mendelian Randomization Study

**DOI:** 10.64898/2026.04.20.26351299

**Authors:** Julia Tomasi, Huilei Xu, Luqing Zhang, Caitlin E Carey, Matthias Schoenberger, Denise P. Yates, JP Casas

## Abstract

**Background:** Elevated lipoprotein(a) [Lp(a)] is a known risk factor for several cardiovascular-related diseases established from multiple genetic and observational studies. However, the underlying mechanisms mediating the effects of Lp(a) levels on cardiovascular disease risk and major adverse cardiovascular events (MACE) are unclear. The aim of this study was to identify proteins downstream of Lp(a) using mendelian randomization (MR) - a genetic causal inference approach.

**Methods:** A two-sample MR was performed by initially identifying Lp(a) genetic instruments based on data from genome wide association studies (GWAS) of Lp(a) blood concentrations. These instruments were then tested for association with proteins from proteomic pQTL data (Olink from UK Biobank, 2940 proteins and SomaScan from deCODE, 4907 proteins).

**Results:** A total of 521 proteins associated with Lp(a) were identified. Using pathway enrichment analysis, the following MACE-relevant pathways were identified comprising a total of 91 Lp(a) downstream proteins: oxidized phospholipid-related, chemotaxis of immune cells and endothelial cell activation, pro-inflammatory monocyte activation, neutrophil activity, coagulation, and lipid metabolism.

**Conclusion:** The results suggest that the influence of Lp(a) treatments is primarily through modifying inflammation rather than lipid-lowering, thus providing insight into the mechanistic framework which mediates the effects of elevated Lp(a) on atherosclerotic cardiovascular disease.

## Introduction

The link between lipoprotein(a) [Lp(a)] levels and adverse cardiovascular (CV) outcomes is well-established. Elevated levels of circulating Lp(a) are recognized as an independent risk factor for atherosclerotic cardiovascular disease (ASCVD) across various stages. In the general population, high Lp(a) levels are associated with an increased risk of incident CV events (Nordestgaard et al., 2010; Tsimikas, 2017). This risk is pronounced in individuals at high risk of ASCVD, such as those with familial hypercholesterolemia or other genetic predispositions (Nordestgaard et al., 2010). Furthermore, in people with established ASCVD, elevated Lp(a) levels are linked to recurrent CV events and the progression of the disease (Boffa & Koschinsky, 2016; Tsimikas, 2017; Welsh et al., 2024). In a recent analysis based on US medical claims between 2012 and 2022 in 273,770 patients with ASCVD, elevated Lp(a) levels were shown to be associated with progressively increasing risk of recurrent ASCVD events irrespective of sex and race/ethnicity (MacDougall et al., 2025). Although the precise mechanisms underlying these properties are not fully understood, some aspects of Lp(a) function remain the subject of ongoing debate (Koschinsky et al., 2024). Traditionally classified as a lipoprotein, Lp(a) appears to have functions that extend beyond merely transporting lipids (Bhatia et al., 2024; Simantiris et al., 2023). Lp(a) is considered to be pro-atherogenic, pro-inflammatory, and pro-thrombotic (Farzam et al., 2026).

Given the paucity of data on the topic, the mechanistic framework underlying the relationship between Lp(a) and ASCVD is poorly understood. The relationship between Lp(a) and ASCVD can be explored using both in vitro or in vivo approaches and human trials. Preclinical models are limited because the *LPA* gene is only expressed in primates, with significant interspecies differences in function and structure. Further, use of in vitro models is limited by the fact that a systemic biological context afforded by in vivo models is missing. Hence, insights obtained from in vitro models need further corroboration from in vivo models. Therefore, analyses using human data as the mode of investigation are likely the best source for understanding the Lp(a)-ASCVD relationship. Most observational studies that described the relationship between Lp(a) levels and cardiovascular diseases lack the level of biological resolution to unpack the mechanisms that mediate the Lp(a) - ASCVD relationship. Currently, antisense and small interfering RNA therapies targeting the *LPA* gene are under investigation, allowing the use of clinical trial data to assess this mechanistic relationship. However, these trials are often limited by small sample sizes from early phase studies (Viney et al., 2016), while data is pending from ongoing cardiovascular outcome trials such as HORIZON (Novartis Pharmaceuticals, 2019).

Mendelian randomization (MR) is an approach which utilizes naturally occurring variants of the *LPA* gene to create a “quasi-randomized” human dataset (Burgess et al., 2019; Sanderson et al., 2022). The random segregation of alleles in *LPA* cis-variants at gamete formation effectively randomizes the dataset, whereby there are alleles in these variants that are linked to elevated Lp(a) levels in genome-wide association studies (GWAS). These variants can then be used to assess whether genetic variants strongly associated with Lp(a) levels are also linked to proteins in the proteome that may affect the risk of ASCVD. This approach enables us to explore potential downstream effects of Lp(a) using a human dataset, overcoming the limitations of other methods.

Genetic evidence suggests a putative causal relationship between Lp(a) levels and ASCVD. Up to 90% of individual variability in Lp(a) levels is due to genetics, so individuals with a family history of ASCVD are highly likely to also have higher levels of Lp(a) (Schmidt et al., 2016; Tsimikas, 2017). Clarke et al. identified two cis genetic variants in the *LPA* gene that explained a significant portion of the variance in Lp(a) levels, with a score created from these variants being associated with higher odds of coronary artery disease (CAD) (Clarke et al., 2009). Burgess et al. found 43 cis-variants that explained a large amount of variance in Lp(a) levels (51-63%) and showed an association with increased coronary heart disease (CHD) risk. Their study indicated that for each 10 mg/dL decrease in Lp(a) levels predicted by their genetic score, there was a 5.8% reduction in CHD risk (Burgess et al., 2018; Schmidt et al., 2016).

The current study employs a MR analysis approach, which aimed to combine causal inference human genetics with proteome data to identify novel proteins related to Lp(a). Upon identification of proteins downstream of Lp(a), we determined whether any were present in biological mechanisms relevant to major adverse cardiovascular events (MACE), thus informing the underlying mechanistic relationship between Lp(a) and ASCVD.

## Methods

### Strategy to understand Lp(a)-MACE mechanisms

This analysis used a two-sample MR approach to identify proteins downstream of Lp(a). Sample 1 consisted of published GWAS data on Lp(a) levels to identify appropriate genetic instruments. To increase robustness of our findings, three GWAS datasets were used: Dron et al., Lamina et al., and Burgess et al (Burgess et al., 2018; Dron et al., 2021; Lamina et al., 2019). These three GWASs had slightly different methods to assay Lp(a), and they all focused on populations of European ancestry. Sample 2 consisted of two published GWASs of proteomics data (i.e., pQTL datasets) that used different technologies to assay the human proteome in peripheral blood. The first was a GWAS of SomaScan proteomics from the deCODE study (Ferkingstad et al., 2021), and the second a GWAS of Olink proteomics from the UK Biobank (Sun et al., 2023). We utilized a two sample MR approach which tested the association between the Lp(a) genetic instruments (from each of the three different sources) with proteins in deCODE (SomaScan) and those in UK Biobank (Olink).

### Selection of Lp(a) instruments

Three GWASs assessing Lp(a) concentrations were employed as sources to select genetic instruments to identify Lp(a) cis-variants as a part of Sample 1. First, genetic variants identified in Burgess et al., (2018) were used as instruments, which included 43 single nucleotide polymorphisms (SNPs). The second was selected from the study by Lamina et al. (2019), which identified a 27 SNP subset from Burgess et al. (2018), with a MAF >= 1%. Lastly, the GWAS of Lp(a) performed by Dron et al., (2021) was used to extract instruments, keeping only the SNPs associated with Lp(a) levels at GWAS significance (p<5e-8) that are independent (r2=0.01) and within the *LPA* gene region (chromosome 6: 160531482-160664275, +/-500kb) (**Table 1**).

**Table 1.** Overview of the three GWASs that investigated genetic associations with Lp(a) concentrations.

|  | Lamina et al. | Burgess et al. | Dron et al. |
| --- | --- | --- | --- |
| **Studies included** | 5 studies (Lp(a) measured the same way in all studies) | 5 studies (Lp(a) not measured the same way in all studies) | 1 – UK Biobank |
| **Sample size** | 13,781 | 48,333 | 321,846 |
| **Assay to measure Lp(a)** | ELISA | Immunoturbidimetric (DiaSys, Technicon Axon, Denka Seiken, or Roche) or ELISA (Innogenetics) | Immunoturbidimetric (Randox Bioscience, UK) |
| **Lp(a) units** | mg/dL | mg/dL | nmol/L (inverse normal transformation) |
| **Number of instruments** | 27 variants | 43 variants | 34 variants |
| **Reason for inclusion** | Unified assay across samples, variants are a subset of Burgess | Most cited | Largest sample with single unified assay |

### Association of Lp(a) with the proteome

The outcome variable was proteome pQTL data. The primary analysis was performed in healthy individuals from the general population, using proteome data from the UK Biobank (Olink, 2940 proteins in 52,363 individuals) and deCODE (SomaScan, 4907 proteins in 35,559 individuals). MR was performed in both datasets separately to assess consistency of results across them. To correct for multiple testing at the proteome level, we used an FDR threshold of 0.01. The primary analysis for two-sample MR was the Inverse Variance weighted (IVW) method (Burgess et al., 2013; Burgess et al., 2019). To assess robustness of significant proteins identified with IVW, we also performed supplemental MR analyses using the weighted median and MR-Egger regression methods.

### Biological interpretation

The results of the MR analysis were used to inform subsequent analyses to interpret the findings. We used several methods to explore pathway enrichment, causality validation, and highlight proteins with the strongest evidence. QIAGEN Ingenuity Pathway Analysis (IPA) and Reactome were used to determine whether the identified significant proteins belong to any biological pathways. To further build support for the protein findings, we assessed whether any significant proteins identified also carry human genetic evidence related to CVD (**Supplementary Table 1**). This evidence included the use of FinnGen’s rare variants tool to identify protein-coding variants (i.e., pLOF variants) from each protein and explore their associations with clinical endpoints related to CHD or stroke, GeneBass to examine the presence of exome-based gene burden results for each protein and associations with CHD/stroke endpoints, and ClinVar and GEL Panel App to determine whether proteins are associated with any CHD/stroke related clinical phenotypes. We also investigated common-variant GWAS evidence, determining whether any significant proteins were also significant in an MR of pQTLs with CHD/stroke, and whether any identified proteins are encoded by a gene with a Locus-to-Gene score ≥0.5 in Open Targets along with association with CHD/stroke terms. The quality (e.g., orthogonal validation) of the SOMAmers/proteins was also extracted.

Primarily using the results from IPA and Reactome for the significant proteins in our MR analyses, six pathways with a known relevance to MACE were identified.

### Strength of evidence assessment

For the MR analyses performed for each Lp(a) instrument, against each pQTL dataset, the absolute beta coefficients were assigned to tertiles to quantify high, medium, or low beta strength. For each of the identified biological pathways, the absolute MR beta values for the proteins in that pathway were averaged. The proportion of proteins that had a high beta strength for each pathway was also calculated.

## Results

### MR: Proteins associated with Lp(a)

The results for the MR analyses using the IVW method against the proteome data with three different Lp(a) instruments (from Burgess et al., Lamina et al., and Dron et al.) are presented in **Supplementary Table 2**. Results from the weighted median and MR-Egger regression methods are presented in **Supplementary Table 3** and **Supplementary Table 4**, respectively. Beta directionality was concordant using weighted median and MR-Egger for all proteins which also showed significant association when assessed with the IVW method. **Supplementary Table 5** annotates the significant IVW MR results, showing the proteomic platform that the significant protein was found in, the specific source for Lp(a) instrument(s) that the protein was significantly associated with, the beta strength for the significant association (high, medium, or low based on tertile assignment), and whether the protein shows additional human genetic evidence (as outlined in the criteria listed in **Supplementary Table 1**). Of the 4907 proteins tested in SomaScan, 232 were significantly associated with at least one of the three tested Lp(a) instrument sources, and of the 2940 proteins tested in Olink, 295 were significant, with a total of 521 unique proteins being significant. Six proteins were significant in both SomaScan and Olink, with only one (PLA2G7) showing concordant beta directionality. All proteins that were significantly associated with Lp(a) in two or more of the tested Lp(a) instrument sources were completely concordant in terms of directionality of the MR beta coefficients, with betas being strongly correlated across all tests (r=0.99). For the number of proteins from SomaScan and Olink that were significant with each of the three tested Lp(a) instrument sources, and how many proteins were significant with multiple, see **Supplementary Figure 1**.

### Mechanisms relevant to MACE associated with proteins downstream of Lp(a)

Based on the 521 significant proteins identified in our MR analyses, six groups of proteins were identified that represent specific pathways potentially related to MACE, with 91 (17%) of the significant proteins being assigned: oxidized phospholipid (OxPL)-related proteins, neutrophil degranulation, pro-inflammatory monocyte activation, chemotaxis immune cells and endothelial cell activation, coagulation/thrombosis, and lipids.

Figure 1 shows the number of proteins in each pathway, the average absolute beta for the proteins in each of the pathways, and the percent of proteins in each pathway that have a high beta strength based on tertile assignment. Notably, the OxPL-related proteins pathway, despite having the fewest number of proteins (n=5), had the greatest proportion of proteins with a high effect size, and the highest average absolute beta overall. Additionally, the pathway with the highest number of proteins was neutrophil degranulation.

The proteins present in each pathway, as well as their function according to the Open Targets Platform and literature, are presented in **Table 2**.

**Figure 1.**
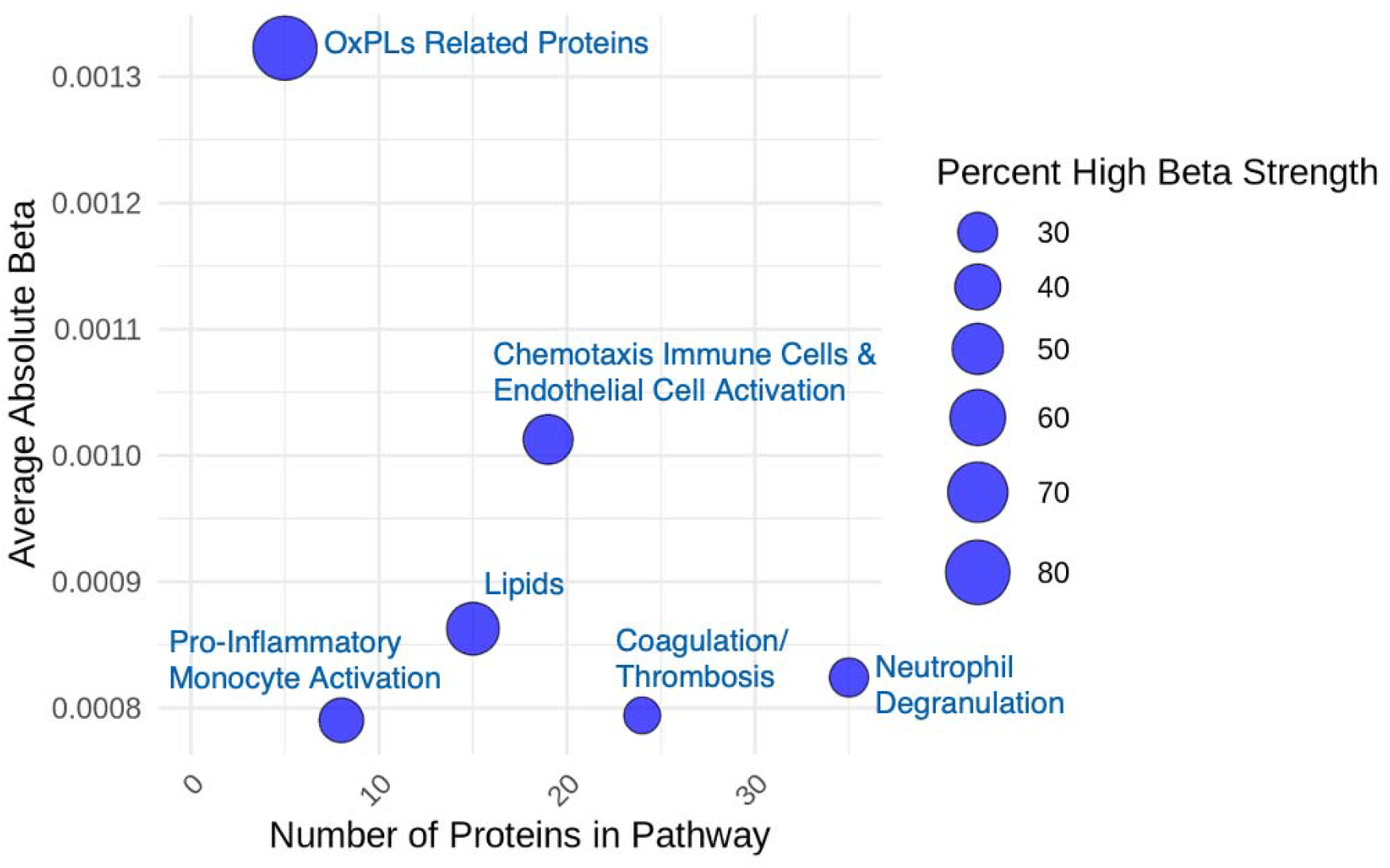
Average absolute beta and strength of beta per pathway OxPL: Oxidized Phospholipids

**Table 2.**
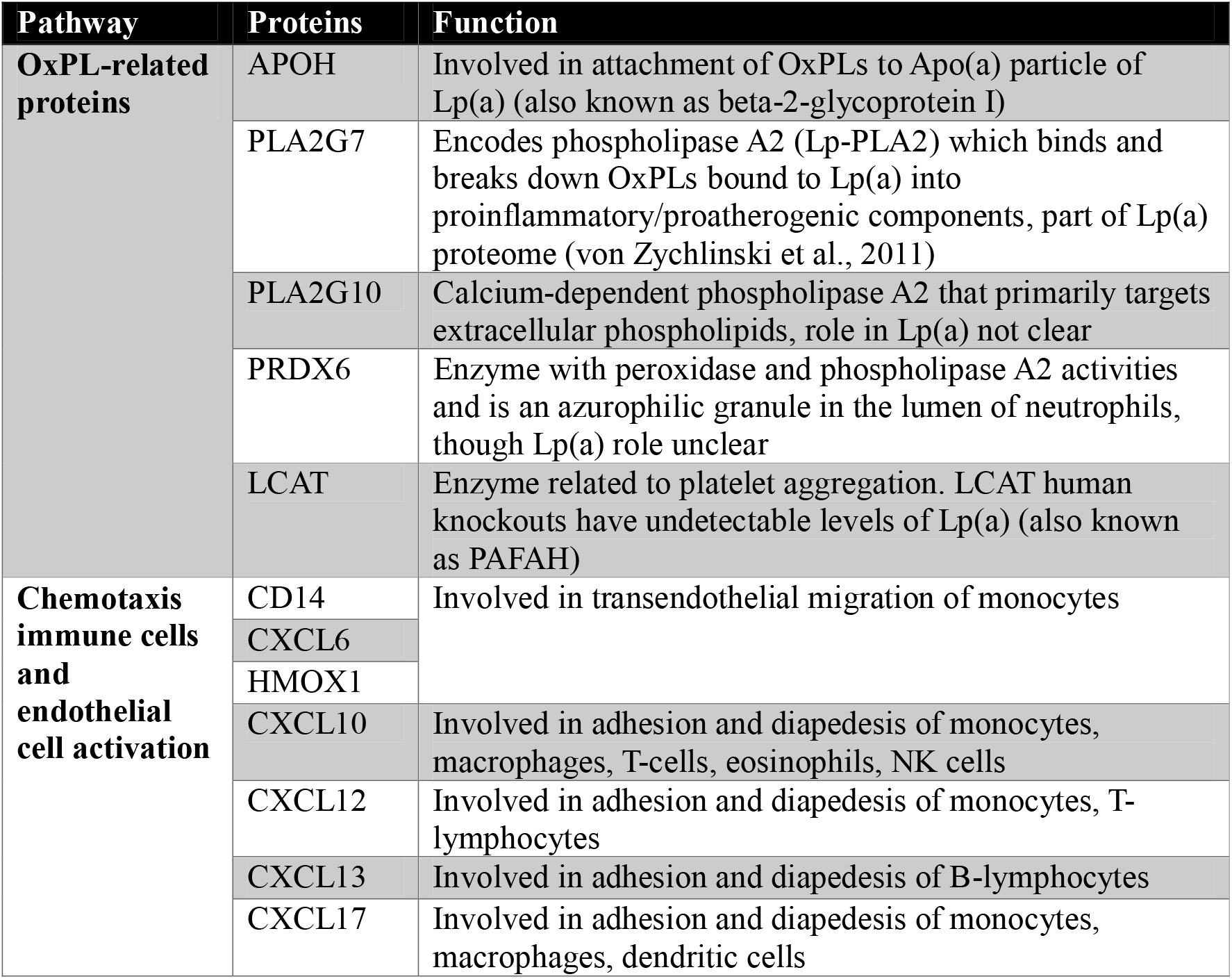
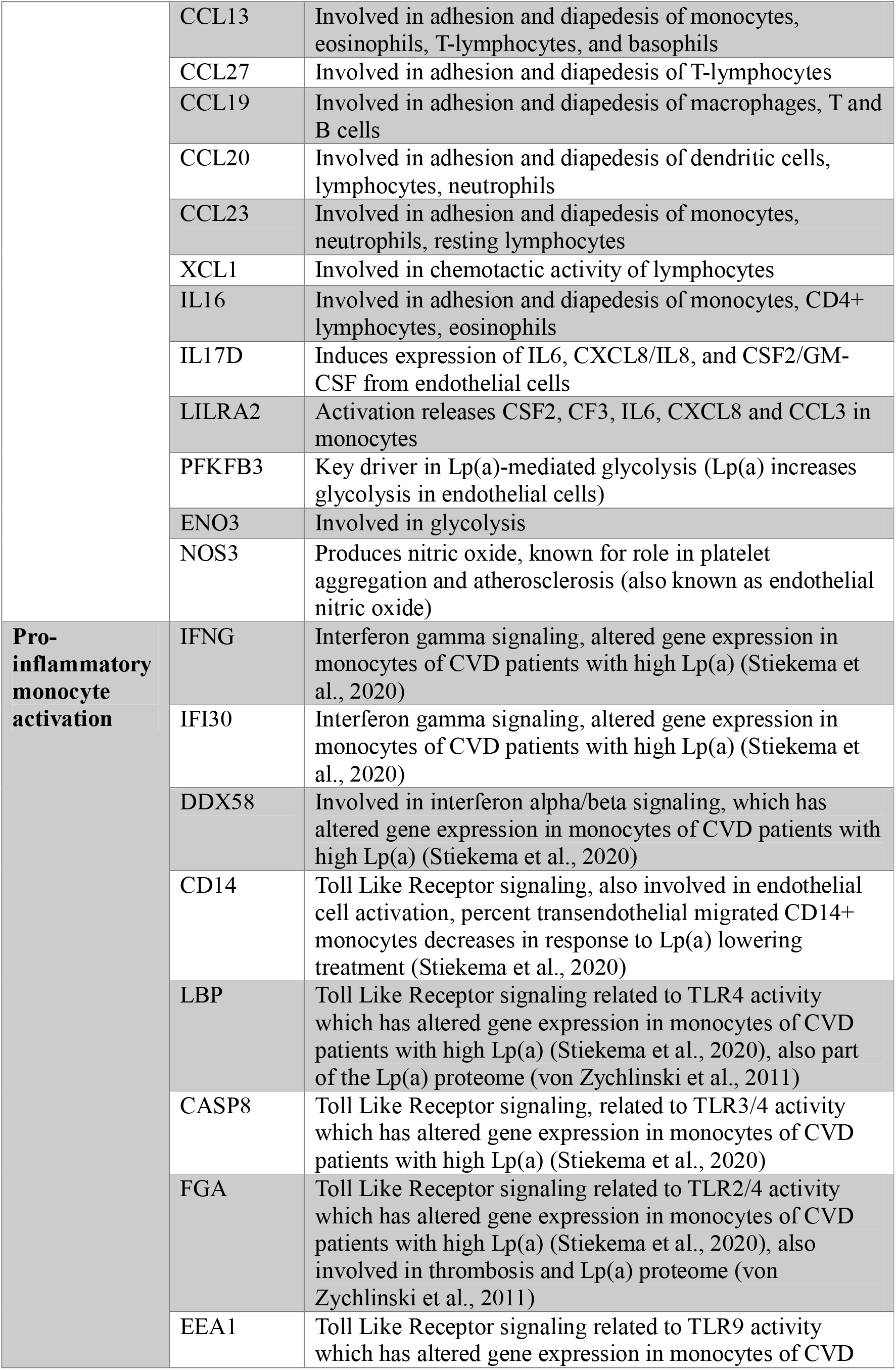
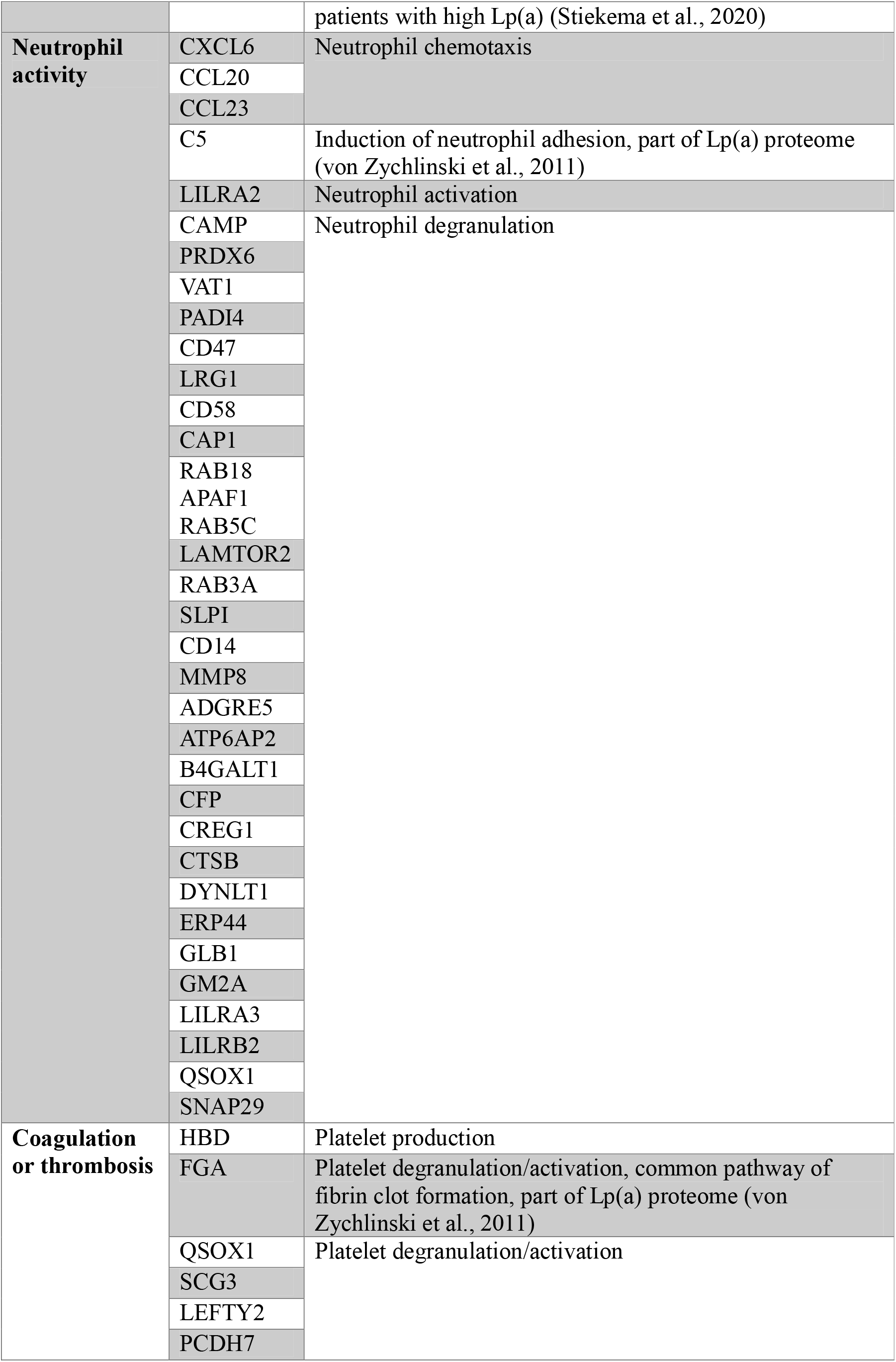
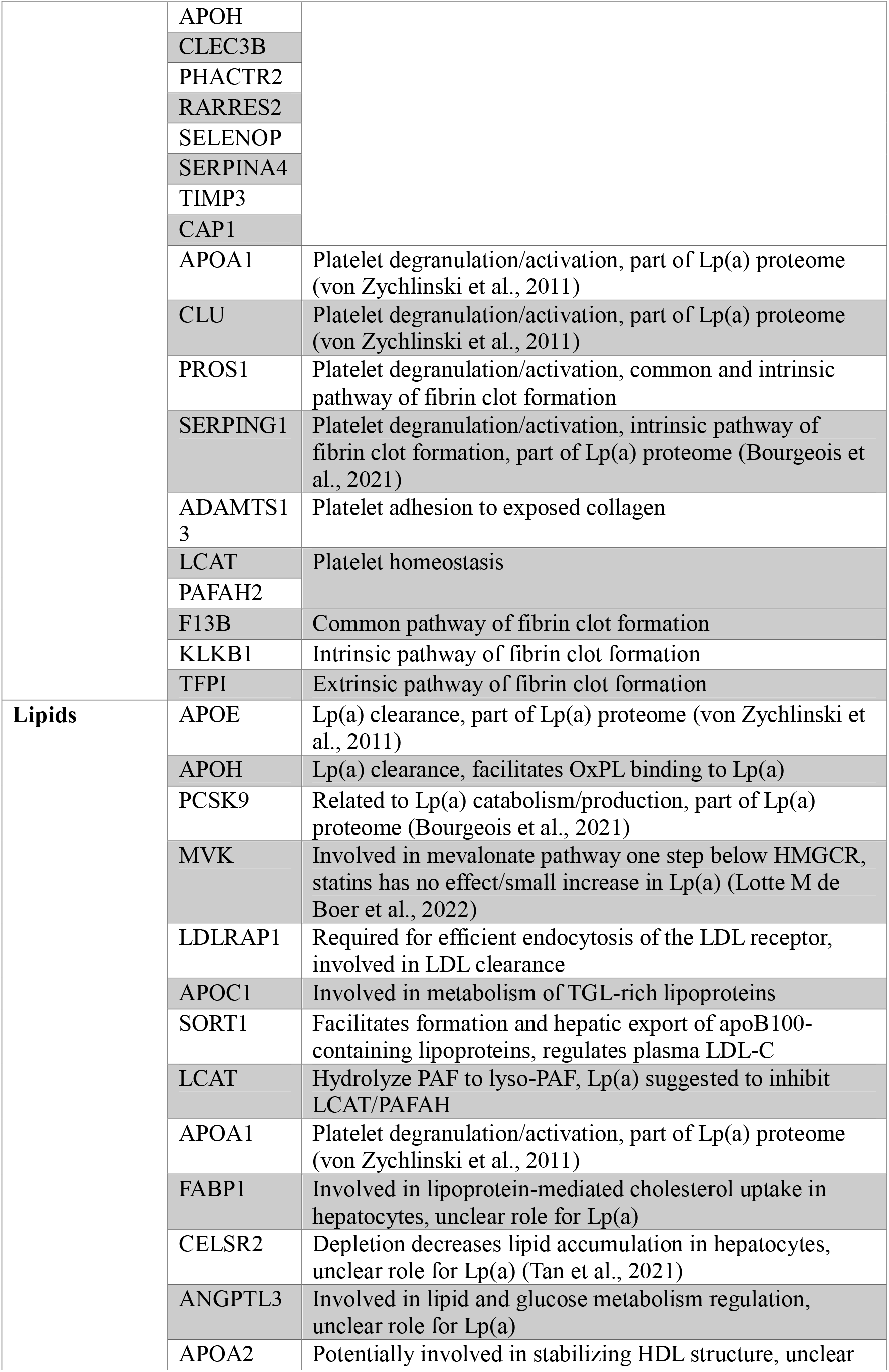
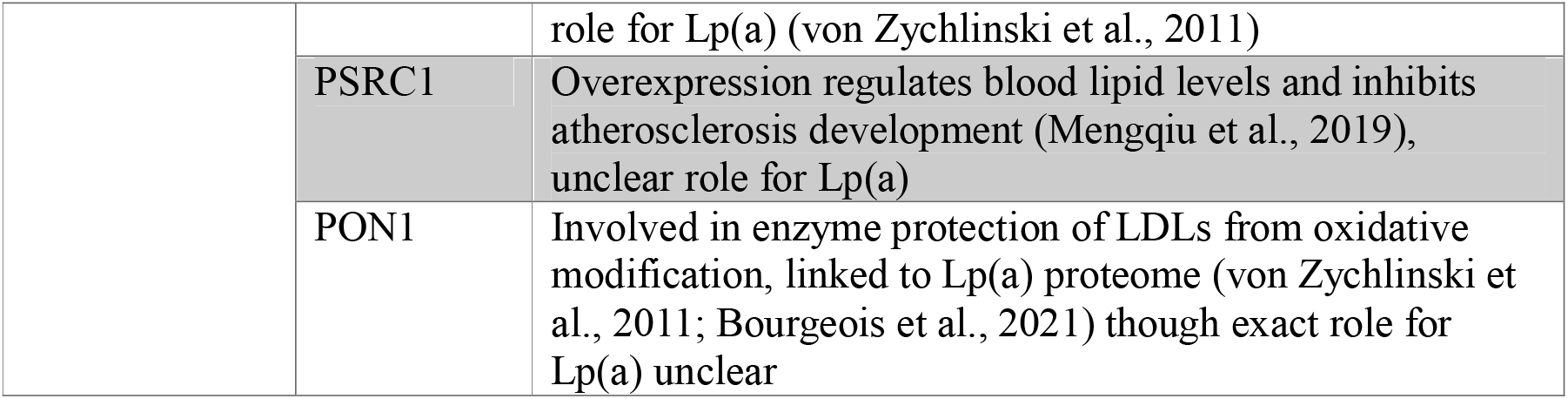
Proteins present in each of the identified pathways and their function.

## Discussion

The current MR analysis used causal inference human genetics with proteome data to identify proteins downstream of Lp(a) to improve understanding of the underlying mechanistic relationships between Lp(a) and cardiovascular disease outcomes. The analysis identified 521 significant proteins associated with Lp(a), with 91 proteins assigned to 6 different biological pathways relevant to MACE. Of these pathways, proteins related to OxPLs showed the strongest effect in the MR analysis for association with Lp(a). Further, the significant proteins found in inflammatory-related paths suggest that monocytes and neutrophils specifically are key peripheral immune cells driving inflammatory processes in relation to Lp(a). Given the abundance of published evidence suggesting that OxPLs are responsible for the inflammatory properties of Lp(a), this OxPL pathway may thus be the key mechanism largely driving other identified mechanisms related to inflammation. Additionally, proteins identified related to lipids have functions relevant to metabolism/catabolism of Lp(a), rather than lipid-elevating properties. Overall, the evidence from the current study suggests that the effects of Lp(a) on MACE outcomes are likely primarily driven by inflammatory effects rather than lipid-lowering.

The results from the current analysis are noteworthy because the MR approach derives data from human genetic and proteomic sources. This approach allows us to minimize bias by leveraging the random assignment of genetic variants at gamete formation to assess whether Lp(a) is causally associated with protein levels across the proteome. Additionally, this analysis used two complementary proteomic databases. Thus, the MR approach taken in the current study provides a more robust assessment of causal relationships to identify proteins downstream of Lp(a) and assign them to relevant biological pathways.

Lp(a) is thought to mediate atherogenesis through mechanisms linked to OxPLs. Lp(a) is the main carrier of OxPLs, with up to 90% of all OxPLs in humans being carried on Lp(a) through apolipoprotein H, or APOH (Tsimikas et al., 2005). This is reflected in the closely positively correlated levels of Lp(a) and OxPLs (Tsimikas et al., 2020). OxPLs originate from the oxidation of polyunsaturated fatty acid residues of phospholipids (Ashraf & Srivastava, 2012). They are created on oxidized low-density lipoprotein (oxLDL), apoptotic cells, oxidized cell membranes, and areas of inflammation (Taleb et al., 2011). Then, OxPLs move from these sources into circulation and are preferentially transferred to and carried by Lp(a) (Tsimikas et al., 2020; Viney et al., 2016). OxPLs are substrates for lipoprotein(a)-associated phospholipase A2 (Lp-PLA2; encoded by PLA2G7), which bind and degrade OxPLs into pro-inflammatory and pro-atherogenic molecules, such as lysophosphatidyl choline (Tsimikas et al., 2005). Such resulting events may contribute to cardiovascular disease, thus suggesting the reduction of Lp(a) levels may remedy this cascade of events. In a phase 2 study where individuals with elevated Lp(a) levels were treated with pelacarsen, an Lp(a)-lowering antisense oligonucleotide, the levels of OxPLs and Lp(a) both showed consistent decreases over time (Viney et al., 2016). Given this mechanism of action, it is expected that CVD patients with elevated Lp(a) may benefit from such a reduction in Lp(a), and subsequently also a reduction in the associated OxPLs with their pro-inflammatory actions, potentially lowering the risk for MACE and other harmful outcomes.

Lp(a) is known to be associated with several pro-inflammatory mechanisms. In the current analysis we identified proteins involved in the activity of monocytes and neutrophils, as well as proteins involved in various other immune cells that are related to the activation of endothelial cells. The effect of Lp(a) on the activation of endothelial cells is thought to be modulated by the ability of Lp(a) to upregulate glycolysis through a key enzyme, 6-phosphofructo-2-kinase/fructose-2,6-biphosphatase 3 (PFKFB3). Chronic endothelial cell activation leads to endothelial cell damage, culminating in endothelial cell dysfunction, thus increasing inflammation and risk for disease (Schnitzler et al., 2020). In a study assessing the effect of stimulating endothelial cells with Lp(a), it was found that such stimulation leads to elevated trans-endothelial migration of monocytes as well as increases in the number of adhered and migrated monocytes (Schnitzler et al., 2020). This is corroborated by Viney et al., who reported decreased trans-endothelial migration of monocytes in response to the reduction in Lp(a) levels by pelacarsen treatment in a phase 2 study (Viney et al., 2016). In another study, the transcriptomics profile in monocytes was investigated before and after Lp(a) lowering through pelacarsen in patients with CVD with elevated Lp(a) (Stiekema et al., 2020). They showed an enrichment of genes related to interferon alpha/beta, interferon gamma, and the toll-like receptor signalling pathway, with expression being lower in controls than patients prior to treatment, while after treatment, the expression decreased in patients moving towards the levels of controls. This effect coincided with a reduction of monocyte trans-endothelial migration following treatment. In our MR analysis, we found proteins present in the pathways that showed differential enrichment in this study, therefore bolstering the evidence for a role of monocytes in Lp(a) function. While much of the evidence from our MR analysis suggests a notable relation of Lp(a) to monocyte activity, we did also find several proteins linked to the adhesion and diapedesis of other immune cells beyond monocytes (e.g., macrophages, T-cells, eosinophils, natural killer cells, etc.).

Proteins related to neutrophil activity were also identified in an abundance in the current analysis. The direct relationship between Lp(a) and neutrophils is unclear with limited evidence available in the literature. However, a study assessing monocyte activation in endothelial cells stimulated with Lp(a) reported the expression of genes related to neutrophils as well, in their transcriptome analysis (Schnitzler et al., 2020). These include proteins related to neutrophil chemotaxis, adhesion, and endothelial migration, suggesting an effect of Lp(a) on neutrophils in its inflammatory cascade in addition to monocytes. The current MR analysis uncovered proteins related to neutrophil function distinct from monocytes, suggesting that the effect of Lp(a) is on both. Further rationale for the involvement of neutrophils to Lp(a) comes from the evidence that OxPLs are not specific to monocytes in their inflammatory activity, and neutrophils are a critical part of the inflammatory cascade in general. Also, neutrophil extracellular traps (NETs), which are created and released following the activation of neutrophils, are found in atherosclerotic lesions (Moschonas & Tselepis, 2019). NETs potentiate the interaction of Lp(a) with platelets to promote aggregatory effects (Bhatia et al., 2024). Therefore, while much of the literature, and the results in the current study, suggest a key downstream effect of monocytes for Lp(a), other immune cells such as neutrophils may carry notable functional implications and therefore deserve further investigation.

The current MR analysis also identified proteins related to coagulation or thrombosis as being linked to Lp(a) downstream effects. Lp(a) is known to bind to platelets with high affinity and activate them, leading to platelet aggregation (Bhatia et al., 2024). Additionally, Lp(a) prevents clot breakdown by inhibiting tissue plasminogen activator (tPA), which prevents plasmin formation and ultimately leads to increased coagulation (Bhatia et al., 2024). However, the effect of Lp(a) on proteins related to coagulation is ambiguous, with contradictory evidence from the literature. GWASs have not shown an association between Lp(a) and deep venous thrombosis (Helgadottir et al., 2012; Kamstrup et al., 2012). This does not exclude a potential arterial effect, however. Further, in clinical trials of pelacarsen so far, an effect on ex vivo plasma clot generation or lysis time has not been observed (Boffa & Koschinsky, 2016). A possible reason for this discrepancy is that plasminogen concentrations are still much higher than Lp(a), and therefore can still exert its effect on clot breakdown despite the presence of Lp(a). Thus, despite the current MR analysis uncovering proteins linked to platelet function and fibrinolysis, it remains unclear if benefits of Lp(a)-lowering therapies are due to coagulation alterations.

The role of elevated lipids in the context of CVD is well established, though the downstream effects of Lp(a) specifically may not be primarily due to lipid elevating mechanisms, as suggested by the lipid-related proteins identified in our MR analysis. In response to Lp(a)-lowering by pelacarsen, LDL-cholesterol change is very minimal, decreasing by 11%, while Lp(a)-cholesterol decreases by 67% (Yeang et al., 2022). This suggests that the effect of such a treatment is specific to Lp(a)-cholesterol and not explained by LDL, unlike other CVD treatments. It is plausible that the enzymes and receptors related to the regulation of LDL-cholesterol also play a role in Lp(a) regulation, though the lipids related to the catabolism of Lp(a) are still relatively unknown and more research is required. Despite their frequent use in CVD and related conditions, the statins have little to no effect on Lp(a) levels (de Boer et al., 2022). The proteins we identified to be associated with Lp(a) that were related to lipid mechanisms appear to function in Lp(a) catabolism or have other non-lipid roles (e.g., platelet function, OxPL binding). However, many proteins we found in this path, despite being lipid-related, have an unclear role on Lp(a) functioning specifically.

These results of the study should be interpreted in light of certain limitations. Firstly, the exploratory nature of the study involved multiple testing. Secondly, there is a lack of tissue specificity, as proteins were measured in the blood, which may not accurately reflect tissue-specific processes. Additionally, there were inconsistencies between Olink and SomaScan, with some proteins being significant in Olink but not in SomaScan, or vice versa. The study also only included data from individuals of European ancestry, necessitating further investigation to determine if the results are applicable to those of other ancestries. Lastly, the three different Lp(a) genetic instrument sources used in the study did not have consistent units or methods of measuring Lp(a). However, it is noteworthy that the significant proteins identified in the MR analyses with these different instruments largely overlapped. For example, 61 of the 62 Olink proteins significant when using the Lp(a) genetic instruments from Dron et al. were also significant when using the instruments from Lamina et al. and Burgess et al.

The current study opens the discussion to further investigations. Similar MR analyses using the metabolome to identify metabolites that may be downstream of Lp(a) will further contextualise the genetic and proteomic results. The results from such an analysis could inform future targets that may influence MACE outcomes. Additionally, testing for clinical relevance is crucial. If data is available, researchers could explore the levels of notable proteins identified in this study in individuals with high Lp(a) or high genetic risk for Lp(a), and determine whether these levels are altered following Lp(a)-reducing treatment. Furthermore, it would be valuable to investigate whether any of these proteins, potentially measured via blood, could predict treatment outcomes for Lp(a)-reducing therapies. Lastly, RNA interference experiments could be conducted to determine the effects of knocking down the expression of genes encoding some notable proteins identified in this study, particularly those related to OxPLs or inflammation, to further uncover resulting mechanistic alterations linked to Lp(a).

## Conclusions

The current study leveraged MR to elucidate the potential proteomic mechanisms downstream of Lp(a) that may underpin the relationship between Lp(a) and MACE. Proteins associated with OxPLs exhibited the most notable effect in relation to Lp(a), supporting prior literature. Monocytes and neutrophils were identified as key peripheral immune cells driving inflammatory processes in relation to Lp(a). Given that OxPLs are known to be the key driver for the inflammatory effects of Lp(a), it is suggested that the OxPLs may be largely promoting these downstream inflammatory mechanisms. Proteins we identified related to lipids appeared to be primarily involved in the metabolism of Lp(a). Our uncovering of coagulation-related proteins associated with Lp(a) maps on to prior literature, though the effect of this pathway remains to be further explored given ambiguous literature suggesting a lack of thrombotic effects of existing Lp(a)-lowering therapies. The results of this study overall suggest that the effect of Lp(a)-lowering treatment may be primarily due to the modification of inflammatory pathways as opposed to lipid-lowering. This approach therefore expands understanding of the mechanisms behind Lp(a) and medications that target it, and how they have evolved from prior treatments used for MACE.

## Data Availability

All data used in the present study are publicly available GWAS summary statistics. Lp(a) GWAS data were obtained from Burgess et al. (2018), Lamina et al. (2019), and Dron et al. (2021). Proteomic pQTL summary statistics were obtained from the deCODE study (Ferkingstad et al., 2021) and the UK Biobank (Sun et al., 2023).

## Data Sharing Statement

All data sources used for the current study are publicly available, from the following studies: Lamina et al., 2019, Burgess et al., 2018, Dron et al., 2021, Sun et al., 2023, Ferkingstad et al., 2021.

## Acknowledgements

The authors acknowledge that Ashwini Kumar K M, M.Sc (Novartis Healthcare Pvt. Ltd., Hyderabad, India), provided medical writing support which was fully funded by Novartis Pharma AG, Basel, Switzerland in accordance with the Good Publication Practice guidelines, 2022.

## Author Contributions

JT contributed to data analysis and interpretation. HX and JP contributed to conception and interpretation. DY supported the interpretation of the work. CEC contributed to the analysis of data. All authors provided input to draft the manuscript or revise. All authors provided a final approval of the version to be published.

