## Supplementary Figure 1 for "Proteomic Insights into Lp(a) Cardiovascular Mechanisms: A Mendelian Randomization Study"

Supplementary Figure 1. Significant association between proteins (from Somascan and Olink) and Lp(a) genetic instruments

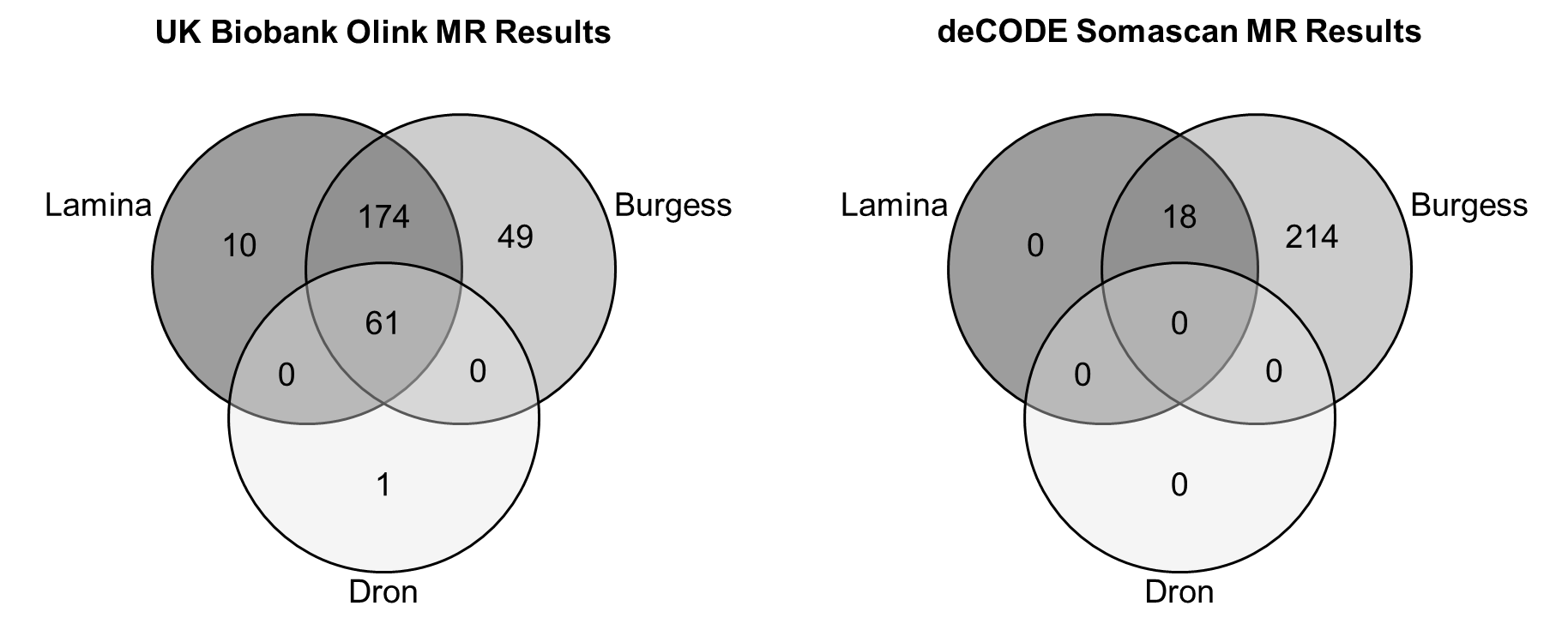
