## Supplementary Table 1 for "Proteomic Insights into Lp(a) Cardiovascular Mechanisms: A Mendelian Randomization Study"

Supplementary Table 1. Human genetic sources to investigate potential causal role of the proteins on MACE components (CHD or stroke)

| Source | Type | Inclusion Criteria and Rationale |
| --- | --- | --- |
| ClinVar | Clinical variants  (protein-coding) | Open Target evidence score ≥0.5 (corresponding to “established risk allele”/”risk factor”/”affects” and above) for CHD or Stroke related terms |
| GEL Panel App | Clinical variants  (protein-coding) | Rating of “amber” (borderline) or “green” (diagnostic-grade). I.e., borderline/Dx for causality on CHD or Stroke related terms |
| FinnGen Protein-Coding Variation | Protein-coding variants | p<5e-5 association with CHD or Stroke related terms |
| GeneBass Gene Burden | Protein-coding variants | p<5e-5 association with CHD or Stroke related terms |
| Open Targets | Common GWAS variants | L2G score ≥0.5: Associated with higher odds of drug target success |
| MR pQTL Analyses | Common GWAS variants | FDR p<0.05 association with CHD (Aragam et al., 2022) or Stroke (Zhou et al., 2022): Associated with 2.4 higher odds of drug-target success |
